# Preliminary High-Level Modelling Estimates of Impacts of Denicotinisation on Smoking Prevalence in Aotearoa New Zealand

**DOI:** 10.1101/2021.08.13.21262035

**Authors:** Nick Wilson, Janet Hoek, Nhung Nghiem, Jennifer Summers, Leah Grout, Richard Edwards

## Abstract

**Aim:** To provide preliminary high-level modelling estimates of the impact of denicotinisation of tobacco on changes in smoking prevalence in Aotearoa New Zealand (NZ).

**Methods:** An Excel spreadsheet was populated with smoking/vaping prevalence data from the NZ Health Survey and business-as-usual trends projected. Using various parameters from the literature (NZ trial data, NZ EASE-ITC Study results), we modelled the impact of denicotinisation of tobacco (with no other tobacco permitted for sale) out to 2025, the year of this country’s Smokefree Goal. Scenario 1 used estimates from a published expert knowledge elicitation process, and Scenario 2 considered the addition of extra mass media campaign and quitline support to the base case.

**Results:** With the denicotinisation intervention, adult daily smoking prevalences were all estimated to decline to under 5% in 2025 for non-Māori and in one scenario for Māori (Indigenous population) (2.5% in Scenario 1). However, prevalence did not fall below five percent in the base case for Māori (7.7%) or with Scenario 2 (5.2%). In the base case, vaping was estimated to increase to 7.9% in the adult population in 2025, and up to 10.7% in one scenario (Scenario 1).

**Conclusions:** This preliminary, high-level modelling suggests a mandated denicotinisation policy for could provide a realistic chance of achieving the NZ Government’s Smokefree 2025 Goal. The probability of success would further increase if supplemented with other interventions such as mass media campaigns with Quitline support (especially if targeted for a predominantly Māori audience). Nevertheless, there is much uncertainty with these preliminary high-level results and more sophisticated modelling is highly desirable.

## INTRODUCTION

Tobacco smoking caused an estimated 4790 premature deaths in Aotearoa New Zealand (NZ) in 2019 alone (95% uncertainty interval [UI]: 4510 to 5100).^1^ The total health loss, including morbidity, in 2019 was 116,000 disability-adjusted life years (DALYs) lost (95%UI: 108,000 to 125,000).^1^ Furthermore, smoking causes health inequities and results in poorer health for Māori (Indigenous population) versus non-Māori.^2 3^ Exposure to second-hand smoke causes a further estimated 347 additional premature deaths per year in New Zealand, and an additional 9022 lost DALYs per year.^4^

This high health burden means that the health benefits of tobacco control can be extremely large. The highest impact intervention in one modelling study (of a sinking lid on tobacco sales) estimated a saving of 1.21 million quality-adjusted life years (QALYs) and NZ$1.71 billion in cost-savings to the health system (lifetime impacts for the population alive in 2011 and undiscounted estimates).^5^ These gains are very large when compared with the majority of health sector interventions in an online league table with hundreds of New Zealand and Australian interventions.^6^ Other likely benefits from enhanced tobacco control include reduced health inequities (as Māori would receive the greater per capita health gain),^5^ and large economic benefits as reduced illness among workers will improve productivity. For example, a New Zealand study reported that “the majority of the health benefit over a 10-year horizon from increasing tobacco taxes is accrued in the working-age population (20-65 years).”^7^

In April 2021, the New Zealand Government published a Discussion Document outlining proposals for an Action Plan to realise the Smokefree Aotearoa 2025 Goal.^8^ This Document has attracted very favourable international attention.^9^ One of the major potential interventions in this Discussion Document was the reduction of nicotine in smoked tobacco products to very low levels (i.e., to non-addictive levels).

Several reviews and commentaries, and many individual studies,^10-35^ have investigated the impact of very low nicotine cigarettes (VLNCs), which are generally defined as having around 0.4 mg or less nicotine per gram tobacco or per cigarette. Overall, this work has concluded that most people who smoke and who are provided with VLNCs find these cigarettes unsatisfying. As a result, study participants often cut down on the number of cigarettes per day, have similar or lower biomarkers of exposure to toxins, experience fewer withdrawal effects, make more quit attempts, and become more likely to quit successfully (see Edwards et al^36^ for a recent review of these issues).

Modelling studies also suggest that a mandated VLNC policy would result in substantial reductions in smoking prevalence and population health gains.^37 38^ A historical modelling study has also estimated that had the tobacco industry introduced VLNCs when the health effects of smoking were established in the 1960s, millions of lives would have been saved.^39^

The VLNC/denicotinisation approach aligns with the findings of a New Zealand Government inquiry by the 2010 Māori Affairs Select Committee, which recommended reducing the additives and nicotine in tobacco to help achieve the proposed Smokefree 2025 Goal (recommendation 9).^40^ This approach is also likely to have public support in New Zealand. For example, 80% of respondents in a recent New Zealand survey of people who smoke, or who have recently quit, supported mandated VLNCs, provided alternative nicotine products were available.^41^ International studies have reported similar very strong support for this policy.^42 43^ For these reasons, we performed high-level modelling work on the likely impact of denicotinisation to inform the New Zealand Government’s upcoming decision-making.

## METHODS

### Base case analysis assumptions

We assumed the following steps and input parameters for our base case (considered most likely) analysis:

1) Consultation and deliberation via parliamentary processes (e.g., Select Committee) on the proposed denicotinisation law was assumed to occur in late 2021. In this year and the next one, the business-as-usual (BAU) downward trends in smoking prevalence for all groups were as per the average trend for the eight year period between 2011/12 and 2019/2020^44^ (the period for which the New Zealand Health Survey [NZHS] was run continuously). For the more recently collected data on vaping in the NZHS, we used the pattern between 2018/19 and 2019/2020 (NZHS data^44^) for the BAU trend.
2) The denicotinisation law was assumed to pass in the year 2022, with an implementation date of 1 March 2023 (i.e., from which point the only tobacco permitted for sale in New Zealand would be denicotinised tobacco).
3) In 2023, and each subsequent year, we assumed that initiation of smoking in the 18-24-year-old age-group would be reduced by 75% (due to the non-addictive nature of the denicotinised tobacco). That meant that each year there would be a reduction in around 6500 smokers (one seventh of the 61,000 smokers in this age-group group multiplied by 75%; NZHS data for 2019/2020^44^). This 75% value is very uncertain but we considered it more realistic than the 50% estimate considered elsewhere.^45^ We did not estimate the proportion of these that would have taken up vaping instead.
4) In 2023, we assumed that 33% of smokers would quit, as per the New Zealand trial data for such denicotinised products (i.e., more specifically, 33% had quit at six months with no reported difference in impact between Māori and non-Māori^30^). The remaining 67% were assumed to continue smoking, using either denicotinised tobacco or regular tobacco (obtained via illicit supply or via home-grown tobacco for personal use, which is legal in New Zealand). Those who quit were assumed to become either quitters or vapers as per the ratios identified in the EASE-ITC Study (preliminary data supplied by the principal investigator Professor Richard Edwards [one of the co-authors of this current study]). Respondents in this study answered the following question: “Which one of the following would you be most likely to do if – the amount of nicotine in cigarettes was greatly reduced so that they are no longer addictive?” Response options included: “quit smoking entirely” (13.5% of respondents, a mix of smokers and recent quitters, gave this answer) and “switch to vaping/ e-cigarettes” (13.2% gave this answer). We assumed no major reductions in the accessibility of vaping products would occur.
5) In the following two years (2024 and 2025) we assumed the same impact as in 2023 (i.e., 33% of smokers using denicotinised tobacco quit per year). We assumed that this relatively high rate of quitting would be sustained due to the non-addictive nature of the denicotinised tobacco product and the growing denormalisation of smoking as additional tobacco control measures described in the Action Plan for Smokefree Aotearoa were implemented.

### Assumptions for the Scenario One Analysis (alternative parameters)

For an alternative approach we considered expert knowledge elicitation work by Apelberg et al,^45^ which has also been used in other modelling work examining denicotinisation in the US.^39^ Apelberg et al gave the following values when using the 50^th^ percentile estimates from the elicitation exercise (with the below averaging the values for male and female smokers):

In the first year, when only denicotinised cigarettes were permitted on the New Zealand market (the year 2023, as per above):

- 50% reduction in initiation (in contrast to the 75% we used in the base case)
- 20% of smokers quit and do not switch products (i.e., end nicotine use completely)
- 37.5% of smokers quit and switch to non-combustible tobacco products (in New Zealand, we assumed these products were only e-cigarettes)

In the second year and each subsequent year up to, and including, 2025, the respective values were:

- 50% reduction in initiation
- 14.3% quit
- 38.3% switch (to vaping as per the first year detailed above)

### Assumptions for the Scenario Two Analysis (extra campaign/Quitline support)

We also considered the impact of adding mass media cessation promotion, with Quitline support, to the base case denicotinisation intervention. The impact of the New Zealand Quitline is well established via multiple studies (including randomised trials) and via a detailed New Zealand modelling study that included media campaign impacts.^46^ We used the results from this previous modelling study to consider the impact of doubling mass media campaign expenditure with Quitline support (a “campaign/service” package). That is, in normal times, the routine campaign/Quitline support (taking Māori men and women combined) accounted for 1.055% of the estimated 4.2% background net cessation rate (a 25.1% contribution [1.055/4.2]) in the 35-54 year old age group (see Table 2 for the 1.055% value and Table A2 for the 4.2% value in the main text and Supplementary file respectively of Nghiem et al^46^). The equivalent proportion from this package for non-Māori was 21.2%. We then applied these two proportions to enhancing the cessation rate associated with denicotinisation. In other words, this extra intervention package was assumed to increase the annual cessation rate from 33% (for denicotinisation as per Walker et al^30^) to 41% for Māori and from 33% to 40% for non-Māori.

The results for the base case and scenario analyses were generated in an Excel spreadsheet, a copy of which is available on request to the authors.

## RESULTS

Estimates for the modelled base case and scenario analyses are detailed in Table 1 and Figures 1 to 3. In the base case model and both scenarios there were major reductions in smoking prevalence for both Māori and non-Māori compared to the BAU projection. If achieving the Smokefree 2025 Goal is assumed to involve adult daily smoking prevalences of under 5%, then Scenario One would achieve the goal for both Māori and non-Māori (prevalences at 2.5% and 0.9% respectively in 2025). However, the base case estimate for Māori at 7.7% in 2025 and Scenario 2 estimate (5.2%) did not realise the Smokefree 2025 Goal. In the base case, vaping was estimated to increase to 7.9% in 2025, and to 10.7% in Scenario One (Table 1, Figure 3).

**Table 1:** **Estimated daily smoking and daily vaping prevalences (%) for business-as-usual (BAU) projection and the base case model and for two scenario analyses as a result of a tobacco denicotinisation policy (in New Zealand adults aged 15+ years, mid-year estimates)**

| Population group | Years up to, and including, the Smokefree Goal year of 2025 |  |  |  |  |  |
| --- | --- | --- | --- | --- | --- | --- |
|  | 2020* | 2021<br>(law debated) | 2022<br>(law passed) | 2023<br>(law comes into force) | 2024<br>(2<sup>nd</sup> year of the law) | 2025<br>(year of the Smokefree 2025 Goal) |
| **BAU** |  |  |  |  |  |  |
| Māori smoking | 28.7 | 27.7 | 26.8 | 25.9 | 25.0 | 24.2 |
| Non-Māori smoking | 10.1 | 9.6 | 9.2 | 8.8 | 8.4 | 8.0 |
| **Base case intervention (denicotinisation)** |  |  |  |  |  |  |
| Māori smoking | 28.7 | 27.7 | 26.8 | 17.7 | 11.7 | 7.7 |
| Non-Māori smoking | 10.1 | 9.6 | 9.2 | 6.1 | 4.0 | 2.7 |
| Total population smoking | 11.1 | 11.1 | 10.6 | 7.0 | 4.6 | 3.1 |
| Total population vaping | 3.5 | 3.8 | 4.2 | 6.0 | 7.1 | 7.9 |
| **Scenario 1 (parameters based on expert elicitation work for the US by Apelberg et al<sup>45</sup>)** |  |  |  |  |  |  |
| Māori smoking | 28.7 | 27.7 | 26.8 | 11.3 | 5.3 | 2.5 |
| Non-Māori smoking | 10.1 | 9.6 | 9.2 | 3.9 | 1.8 | 0.9 |
| Total population smoking | 11.1 | 11.1 | 10.6 | 4.5 | 2.1 | 1.0 |
| Total population vaping | 3.5 | 3.8 | 4.2 | 8.2 | 9.9 | 10.7 |
| **Scenario 2 (adding to the base case by doubling mass media campaign/Quitline support)** |  |  |  |  |  |  |
| Māori smoking | 28.7 | 27.7 | 26.8 | 15.5 | 9.0 | 5.2 |
| Non-Māori smoking | 10.1 | 9.6 | 9.2 | 5.4 | 3.2 | 1.9 |
| Total population smoking | 11.1 | 11.1 | 10.6 | 6.3 | 3.7 | 2.2 |
| Total population vaping | 3.5 | 3.8 | 4.2 | 6.0 | 7.0 | 7.6 |

\* New Zealand Health Survey data for 2019-2020.<sup>44</sup> Technically for the 2020 year these data were collected during 2019/2020 year with some data collection limited by the COVID-19 pandemic in early 2020. Also this survey uses the term “European/Other” while we have used the broadly similar: “non-Māori”.

**Figure 1:**
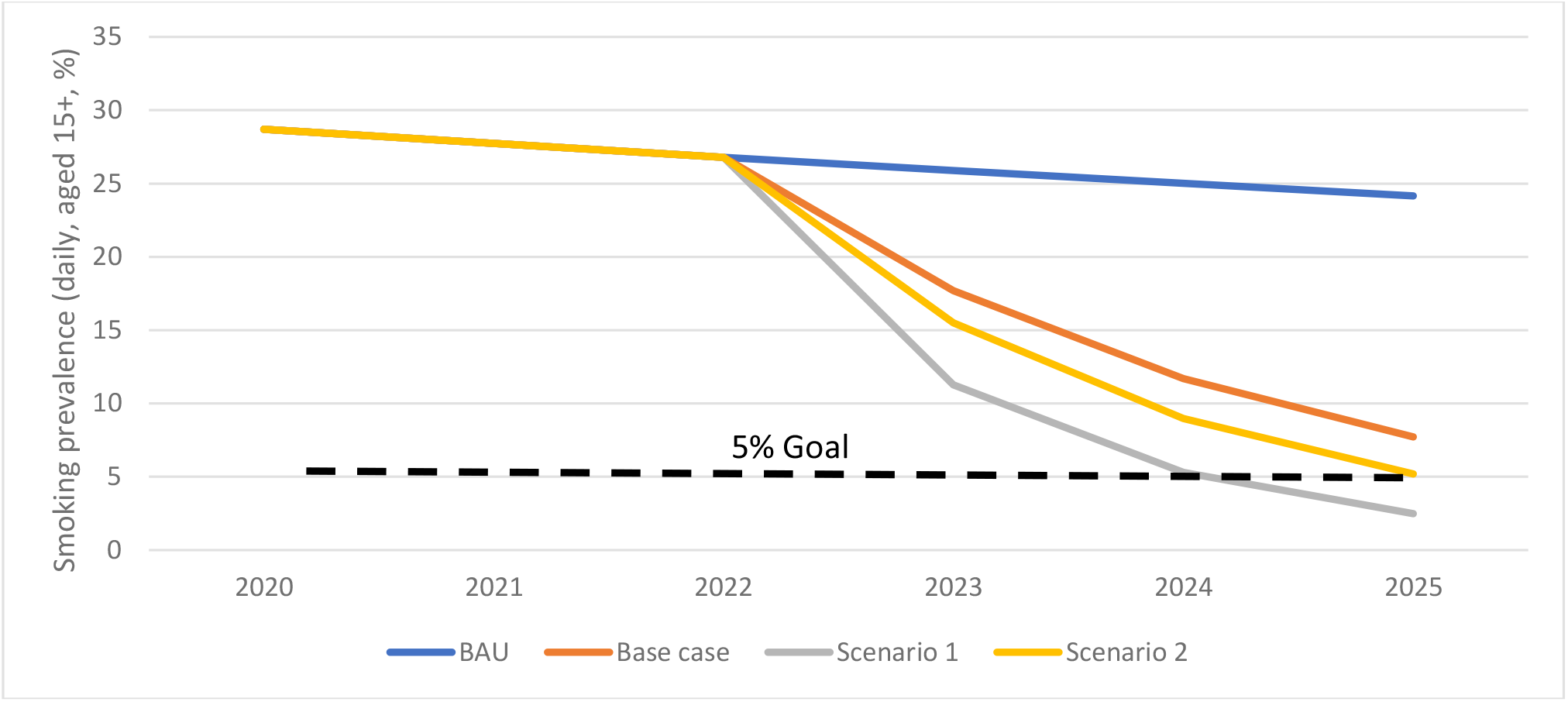
Estimated daily smoking prevalence among Māori for the BAU projection and as a result of a tobacco denicotinisation policy (as per data in Table 1)

**Figure 2:**
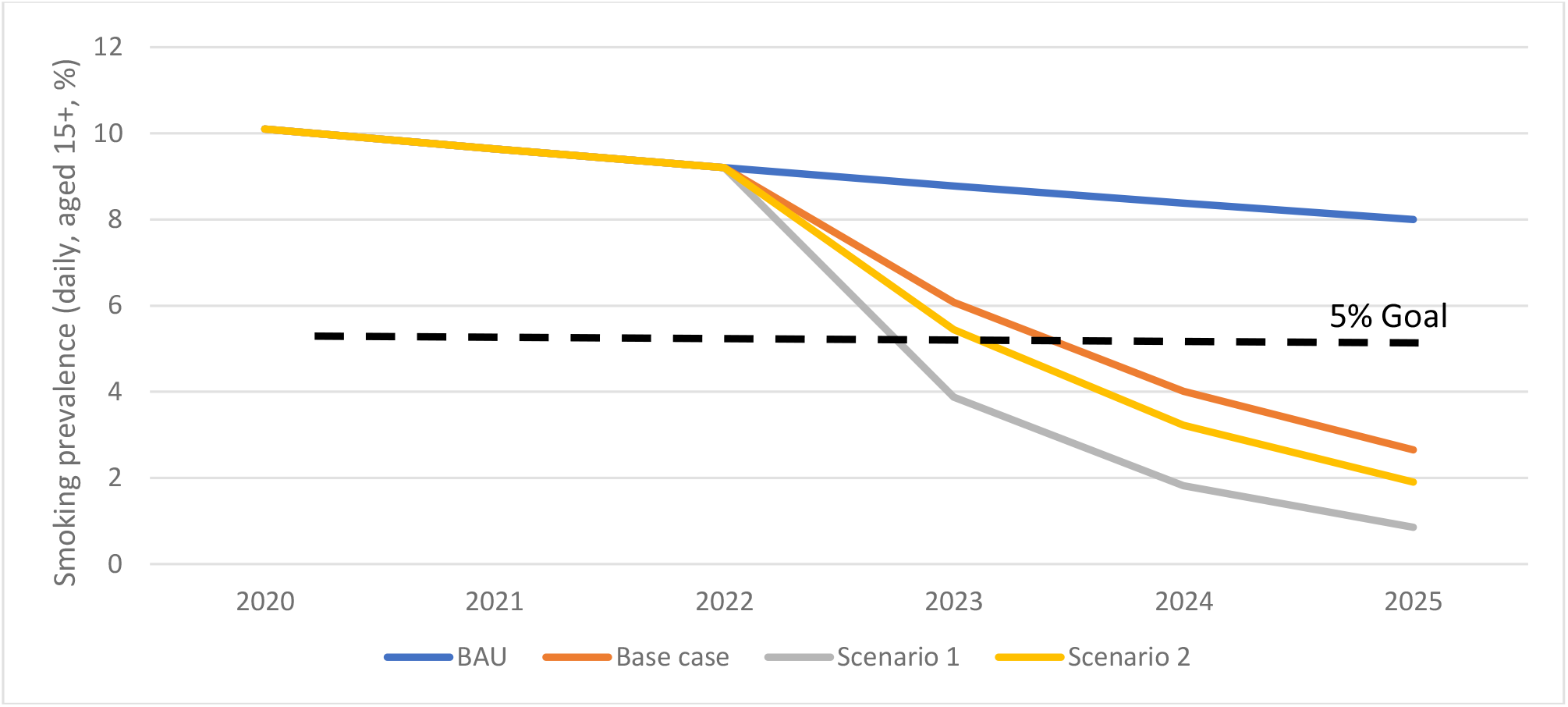
Estimated daily smoking prevalence among non-Māori in the BAU projection and as a result of a tobacco denicotinisation policy (as per data in Table 1)

**Figure 3:**
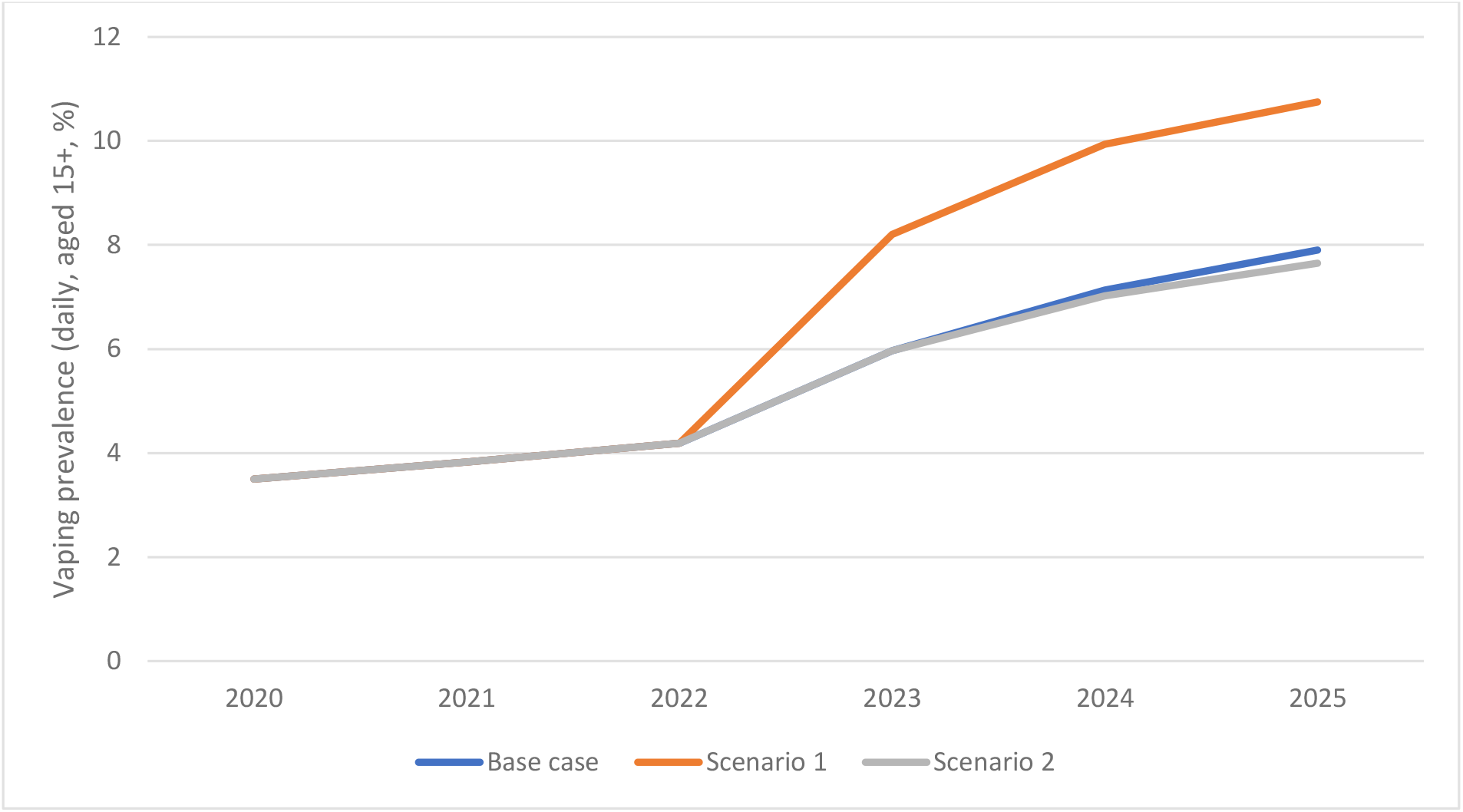
Estimated vaping prevalence in the total population for the BAU projection and as a result of a tobacco denicotinisation policy (as per data in Table 1)

## DISCUSSION

These preliminary high-level results suggest that a tobacco denicotinisation law could come close to achieving, or may potentially achieve, the New Zealand Government’s Smokefree 2025 Goal. However, to be more certain about achieving the goal for Māori, denicotinisation would probably need to supplemented with media campaigns and enhanced Quitline support that goes beyond the doubling of the current level used in Scenario 2. Targeting these campaigns to Māori audiences could build on the success of “by Māori for Māori” campaigns in the past (e.g., the “It’s About Whanau” campaign^47 48^). The addition of other complementary strategies (as outlined in the Discussion Document^8^) could also increase the likelihood that smoking prevalence will fall below 5% among Māori.

However, a partial consequence to these potential outcomes following denicotinisation would probably be a rise in vaping prevalence (as per Figure 3). Vaping still typically involves nicotine addiction, ongoing costs to users, and potential long-term harms to health which are likely to be higher than previously thought.^49^ Nevertheless, our estimates of vaping prevalence may be over-stated if ex-smokers who vape subsequently quit vaping at higher than levels seen to date. But on the other hand, our estimates do not consider any increased vaping uptake among those youth who do not initiate smoking because denicotinised tobacco is non-addictive.

The uncertainty with these high-level modelling results needs to be emphasised, given incomplete international experience on the effects of denicotinising a country’s entire tobacco supply. Therefore, we recommend that these results are considered preliminary until New Zealand Government agencies can commission more detailed analyses (e.g., similar to the much more elaborate tobacco control modelling exercises in New Zealand^7 46 50-54^). Such modelling can capture more epidemiologically precise details of uncertainty intervals, and quantify impacts on quality-adjusted life-years saved, health inequities, and savings in health costs.

When considering uncertainty with these results, it is important to note that the 33% quit rate used in the base case after denicotinisation is introduced may be conservative (the 33% value is based on the New Zealand trial by Walker et al^30^). This particular trial was undertaken in a BAU New Zealand context, where participants could easily access regular tobacco from thousands of retail outlets and via social sources, such as friends and family members. Furthermore, vaping products were not as widely available when this trial was undertaken and thus were not the viable alternative that they are today. If only denicotinised tobacco was available, the only legal alternatives to this would be quitting, adopting vaping, or using pharmaceutical grade products (e.g., nicotine gum and patches). On the other hand, we have assumed that among the 33% of people estimated to quit, none relapse; however, it is possible that some would relapse and use either illicit or home-grown tobacco.

Indeed, another limitation is uncertainty about the size of the illicit and home-grown markets following a denicotinisation law coming into force. In terms of the current size of the illicit market, reviews have noted the limited number of independent (non-tobacco industry funded) studies for New Zealand.^55^ Nevertheless, the most recent independent estimate from 2013 was that illicit products made up only 1.8-3.8% of the New Zealand market.^56^ Commentators have also suggested that any increase in illicit trade is likely to be modest and would not undermine the substantial positive effects of a denicotinisation policy in reducing smoking prevalence.^57^ Furthermore, New Zealand has very strong border controls and surveillance which, coupled with its remote island status, reduce the likelihood that smuggled tobacco would become a major problem (at least compared to European countries). Nonetheless, surveillance and enforcement should ideally be strengthened further during this period (as the Government’s Discussion Document^8^ suggests).

Regarding home-grown tobacco, it also seems likely that supply via this source would not be large, given the difficulties of growing and curing tobacco within New Zealand (e.g., owing to high humidity in much of the country). Before growing commercial tobacco crops ended in New Zealand, efforts to grow tobacco outside the Nelson-Motueka area were not particularly successful. The “roughness” of home-grown product (lacking flavours and additives such as humectants), may also not suit the taste of most New Zealand smokers, especially compared to vaping. Concern with toxins from mould growth on home-grown tobacco product could also be a potential theme in Ministry of Health communications to smokers when the new policy is enacted. The government could also reduce the amount of tobacco that may be legally grown for personal use by home-growers or even require home-growers to have a licence to grow (to allow for occasional spot checks and compliance with the law).

## CONCLUSIONS

This preliminary, high-level modelling suggests a mandated denicotinisation policy could provide a realistic chance of achieving the New Zealand Government’s Smokefree 2025 Goal. The probability of success would further increase if supplemented with other interventions such as mass media campaigns with Quitline support (especially if predominantly for a Māori audience). Nevertheless, there is much uncertainty with these preliminary high-level results and more sophisticated modelling is highly desirable to reduce uncertainty; quantify impacts on quality-adjusted life-years saved and health inequities, and estimate savings in health costs.

## Data Availability

The Excel file with the data and modelling results is available on request of the authors.

## Acknowledgements

The authors thank the Health Research Council of NZ (Grant 10/248) for support with developing the BODE^3^ tobacco control model, which has helped provide relevant background parameters for the work in this particular report.

## References

1. GBD Tobacco Collaborators. Spatial, temporal, and demographic patterns in prevalence of smoking tobacco use and attributable disease burden in 204 countries and territories, 1990-2019: a systematic analysis from the Global Burden of Disease Study 2019. Lancet 2021;(E-publication 31 May).

2. Blakely T, Disney G, Valeri L, Atkinson J, Teng A, Wilson N, Gurrin L. Socioeconomic and Tobacco Mediation of Ethnic Inequalities in Mortality over Time: Repeated Census-mortality Cohort Studies, 1981 to 2011. Epidemiology 2018;29:506–16.

3. Teng A, Blakely T, Atkinson J, Kalediene R, Leinsalu M, Martikainen PT, Rychtarikova J, Mackenbach JP. Changing social inequalities in smoking, obesity and cause-specific mortality: Cross-national comparisons using compass typology. PLoS One 2020;15:e0232971.

4. Institute of Health Metrics and Evaluation. GHDx (Global Health Data Exchange); Results for New Zealand (on 24 May 2021). Seattle; Institute of Health Metrics and Evaluation. http://ghdx.healthdata.org/gbd-results-tool.

5. van der Deen FS, Wilson N, Cleghorn CL, Kvizhinadze G, Cobiac LJ, Nghiem N, Blakely T. Impact of five tobacco endgame strategies on future smoking prevalence, population health and health system costs: two modelling studies to inform the tobacco endgame. Tob Control 2018;27:278–86.

6. University of Otago & University of Melbourne. ANZ-HILT: Australia and New Zealand Health Intervention League Table (Vers 2.0) 2019 [Available from: https://league-table.shinyapps.io/bode3/].

7. Cleghorn CL, Blakely T, Kvizhinadze G, van der Deen FS, Nghiem N, Cobiac LJ, Wilson N. Impact of increasing tobacco taxes on working-age adults: short-term health gain, health equity and cost savings. Tob Control 2018;27:e167–e70.

8. Ministry of Health. Proposals for a Smokefree Aotearoa 2025 Action Plan: Discussion document. Wellington: Ministry of Health 2021. https://www.health.govt.nz/system/files/documents/publications/proposals_for_a_smokefree_aotearoa_2025_action_plan-final.pdf

9. Daube M, Maddox R. Impossible until implemented: New Zealand shows the way. Tob Control 2021.

10. Benowitz NL, Donny EC, Hatsukami DK. Reduced nicotine content cigarettes, e - cigarettes and the cigarette end game. Addiction 2017;112:6–7.

11. Benowitz NL, Henningfield JE. Reducing the nicotine content to make cigarettes less addictive. Tob Control 2013;22 Suppl 1:i14–7.

12. Benowitz NL, Henningfield JE. Nicotine Reduction Strategy: State of the science and challenges to tobacco control policy and FDA tobacco product regulation. Prev Med 2018;117:5–7.

13. Donny EC, Walker N, Hatsukami D, Bullen C. Reducing the nicotine content of combusted tobacco products sold in New Zealand. Tob Control 2017:e37–e42.

14. Gottleib S, Zeller M. A nicotine-focused framework for public health. N Engl J Med 2017;377:1111–14.

15. Hatsukam DK, Kotlyar1 M, Hertsgaard LA, Zhang Y, Carmella SG, Jensen JA, Allen SS, Shield PG, Murphy SE, Stepanov I, Hecht1 SS. Reduced nicotine content cigarettes: effects on toxicant exposure, dependence and cessation. Addiction 2010;105:343–55.

16. Hatsukami DK, Perkins KA, Lesage MG, Ashley DL, Henningfield JE, Benowitz NL, Backinger CL, Zeller M. Nicotine reduction revisited: science and future directions. Tob Control 2010;19:e1–10.

17. World Health Organization Study Group on Tobacco Regulaton. Report on the scientific basis of tobacco product regulation. Seventh report of a WHO Study Group. Geneva: World Health Organization 2019.

18. Benowitz NL, Dains KM, Hall SM, Stewart S, Wilson M, Dempsey D, Jacob P, 3rd. Smoking behavior and exposure to tobacco toxicants during 6 months of smoking progressively reduced nicotine content cigarettes. Cancer Epidemiol Biomarkers Prev 2012;21:761–9.

19. Benowitz NL, Hall SM, Stewart S, Wilson M, Dempsey D, Jacob P, 3rd. Nicotine and carcinogen exposure with smoking of progressively reduced nicotine content cigarette. Cancer Epidemiol Biomarkers Prev 2007;16:2479–85.

20. Dermody SS, Donny EC, Hertsgaard LA, Hatsukami DK. Greater reductions in nicotine exposure while smoking very low nicotine content cigarettes predict smoking cessation. Tob Control 2015;24:536–9.

21. Ding YS, Ward J, Hammond D, Watson CH. Mouth-level intake of benzo[a]pyrene from reduced nicotine cigarettes. Int J Environ Res Public Health 2014;11:11898–914.

22. Donny EC, Denlinger RL, Tidey JW, Koopmeiners JS, Benowitz NL, Vandrey RG, al’Absi M, Carmella SG, Cinciripini PM, Dermody SS, Drobes DJ, Hecht SS, Jensen J, Lane T, Le CT, McClernon FJ, Montoya ID, Murphy SE, Robinson JD, Stitzer ML, Strasser AA, Tindle H, Hatsukami DK. Randomized Trial of Reduced-Nicotine Standards for Cigarettes. N Engl J Med 2015;373:1340–9.

23. Donny EC, Jones M. Prolonged exposure to denicotinized cigarettes with or without transdermal nicotine. Drug Alcohol Depend 2009;104:23–33.

24. Hatsukami D.K., Luo X., Jensen J.A., M. aA, Allen S.S., al e. Effect of Immediate vs Gradual Reduction in Nicotine Content of Cigarettes on Biomarkers of Smoke Exposure: A Randomized Clinical Trial. JAMA 2018;320.

25. Hatsukami DK, Kotlyar M, Hertsgaard LA, Zhang Y, Carmella SG, Jensen JA, Allen SS, Shields PG, Murphy SE, Stepanov I, Hecht SS. Reduced nicotine content cigarettes: effects on toxicant exposure, dependence and cessation. Addiction 2010;105:343–55.

26. Hatsukami DK, Luo X, Dick L, Kangkum M, Allen SS, Murphy SE, Hecht SS, Shields PG, al’Absi M. Reduced nicotine content cigarettes and use of alternative nicotine products: exploratory trial. Addiction 2017;112:156–67.

27. McRobbie H, Przulj D, Smith KM, Cornwall D. Complementing the Standard Multicomponent Treatment for Smokers With Denicotinized Cigarettes: A Randomized Trial. Nicotine Tob Res 2015;18:1134–41.

28. Smith TT, Koopmeiners JS, White CM, Denlinger-Apte RL, Pacek LR, De Jesus VR, Wang L, Watson C, Blount BC, Hatsukami DK, Benowitz NL, Donny EC, Carpenter MJ. The Impact of Exclusive Use of Very Low Nicotine Cigarettes on Compensatory Smoking: An Inpatient Crossover Clinical Trial. Cancer Epidemiol Biomarkers Prev 2020;29:880–86.

29. Walker N, Fraser T, Howe C, Laugesen M, Truman P, Parag V, Glover M, Bullen C. Abrupt nicotine reduction as an endgame policy: a randomised trial. Tob Control 2014.

30. Walker N, Howe C, Bullen C, Grigg M, Glover M, McRobbie H, Laugesen M, Parag V, Whittaker R. The combined effect of very low nicotine content cigarettes, used as an adjunct to usual Quitline care (nicotine replacement therapy and behavioural support), on smoking cessation: a randomized controlled trial. Addiction 2012;107:1857–67.

31. Hammond D, O’Connor RJ. Reduced nicotine cigarettes: smoking behavior and biomarkers of exposure among smokers not intending to quit. Cancer Epidemiol Biomarkers Prev 2014;23:2032–40.

32. Benowitz NL, Jacob P, 3rd, Herrera B. Nicotine intake and dose response when smoking reduced-nicotine content cigarettes. Clin Pharmacol Ther 2006;80:703–14.

33. Benowitz NL, Nardone N, Dains KM, Hall SM, Stewart S, Dempsey D, Jacob P, 3rd. Effect of reducing the nicotine content of cigarettes on cigarette smoking behavior and tobacco smoke toxicant exposure: 2-year follow up. Addiction 2015;110:1667–75.

34. Mercincavage M, Souprountchouk V, Tang KZ, Dumont RL, Wileyto EP, Carmella SG, Hecht SS, Strasser AA. A Randomized Controlled Trial of Progressively Reduced Nicotine Content Cigarettes on Smoking Behaviors, Biomarkers of Exposure, and Subjective Ratings. Cancer Epidemiol Biomarkers Prev 2016;25:1125–33.

35. Smith TT, Koopmeiners JS, Tessier KM, Davis EM, Conklin CA, Denlinger-Apte RL, Lane T, Murphy SE, Tidey JW, Hatsukami DK, Donny EC. Randomized Trial of Low-Nicotine Cigarettes and Transdermal Nicotine. Am J Prev Med 2019;57:515–24.

36. Edwards R, Hoek J, Wilson N, Bullen C. Reducing nicotine in smoked tobacco products: A pivotal feature of the proposals for achieving Smokefree Aotearoa 2025. Public Health Expert 2021;(30 April). https://blogs.otago.ac.nz/pubhealthexpert/reducing-nicotine-in-smoked-tobacco-products-a-pivotal-feature-of-the-proposals-for-achieving-smokefree-aotearoa-2025/.

37. Apelberg BJ, Feirman SP, Salazar E, Corey CG, Ambrose BK, Paredes A, Richman E, Verzi SJ, Vugrin ED, Brodsky NS, Rostron BL. Potential Public Health Effects of Reducing Nicotine Levels in Cigarettes in the United States. N Engl J Med 2018;378:1725–33.

38. Tengs TO, Ahmad S, Savage JM, Moore R, Gage E. The AMA proposal to mandate nicotine reduction in cigarettes: a simulation of the population health impacts. Prev Med 2005;40:170–80.

39. Levy DT, Cummings KM, Heckman BW, Li Y, Yuan Z, Smith TT, Meza R. The Public Health Gains Had Cigarette Companies Chosen to Sell Very Low Nicotine Cigarettes. Nicotine Tob Res 2021;23:438–46.

40. New Zealand Parliament. Inquiry into the tobacco industry in Aotearoa and the consequences of tobacco use for Māori. Report of the Māori Affairs Select Committee. Wellington: New Zealand Parliament, 2010.

41. McKiernan A, Stanley J, Waa AM, Kaai SC, Quah ACK, Fong GT, Edwards R. Beliefs among Adult Smokers and Quitters about Nicotine and Denicotinized Cigarettes in the 2016-17 ITC New Zealand Survey. Tob Regul Sci 2019;5:400–09.

42. Chung-Hall J, Fong GT, Driezen P, Craig L. Smokers’ support for tobacco endgame measures in Canada: findings from the 2016 International Tobacco Control Smoking and Vaping Survey. CMAJ Open 2018;6:E412–E22.

43. Bolcic-Jankovic D, Biener L. Public opinion about FDA regulation of menthol and nicotine. Tob Control 2015;24:e241–5.

44. Ministry of Health. New Zealand Health Survey [2019-2020 data]. https://minhealthnz.shinyapps.io/nz-health-survey-2019-20-annual-data-explorer/_w_f5df888f/#!/explore-indicators.

45. Apelberg BJ, Feirman SP, Salazar E, Corey CG, Ambrose BK, Paredes A, Richman E, Verzi SJ, Vugrin ED, Brodsky NS, Rostron BL. Potential Public Health Effects of Reducing Nicotine Levels in Cigarettes in the United States. N Engl J Med 2018;378:1725–33.

46. Nghiem N, Cleghorn CL, Leung W, Nair N, van der Deen FS, Blakely T, Wilson N. A national quitline service and its promotion in the mass media: modelling the health gain, health equity and cost-utility. Tob Control 2018;27:434–41.

47. Wilson N, Grigg M, Graham L, Cameron G. The effectiveness of television advertising campaigns on generating calls to a national Quitline by Maori. Tob Control 2005;14:284–6.

48. Grigg M, Waa A, Bradbrook SK. Response to an indigenous smoking cessation media campaign-it’s about whanau. Aust N Z J Public Health 2008;32:559–64.

49. Wilson N, Summers J, Ait Ouakrim D, Hoek J, Edwards R, Blakely T. Improving on estimates of the potential relative harm to health from using modern ENDS (vaping) compared to tobacco smoking. medRxiv 2021;(27 June). https://www.medrxiv.org/content/10.1101/2020.12.22.20248737v2.

50. Blakely T, Cobiac LJ, Cleghorn CL, Pearson AL, van der Deen FS, Kvizhinadze G, Nghiem N, McLeod M, Wilson N. Health, health inequality, and cos t impacts of annual increases in tobacco tax: Multistate life table modeling in New Zealand. PLoS Med 2015;12:e1001856.

51. Petrović-van der Deen FS, Blakely T, Kvizhinadze G, Cleghorn CL, Cobiac LJ, Wilson N. Restricting tobacco sales to only pharmacies combined with cessation advice: a modelling study of the future smoking prevalence, health and cost impacts. Tob Control 2019;28:643–50.

52. van der Deen FS, Wilson N, Cleghorn CL, Kvizhinadze G, Cobiac LJ, Nghiem N, Blakely T. Impact of five tobacco endgame strategies on future smoking prevalence, population health and health system costs: two modelling studies to inform the tobacco endgame. Tob Control 2018;27:278–86.

53. Pearson AL, Cleghorn CL, van der Deen FS, Cobiac LJ, Kvizhinadze G, Nghiem N, Blakely T, Wilson N. Tobacco retail outlet restrictions: health and cost impacts from multistate life-table modelling in a national population. Tob Control 2017;26:579–85.

54. Nghiem N, Leung W, Cleghorn C, Blakely T, Wilson N. Mass media promotion of a smartphone smoking cessation app: modelled health and cost-saving impacts. BMC Public Health 2019;19:283.

55. Ernst and Young. Evaluation of the tobacco excise increases – Final Report – 27 November 2018. Wellington: Ministry of Health, 2018.

56. Ajmal A, Veng Ian U. Tobacco tax and the illicit trade in tobacco products in New Zealand. Aust N Z J Public Health 2015;39:116–20.

57. Lindblom EN. Illicit Trade Poses No Threat to an FDA Rule to Minimize Nicotine in Smoked Tobacco Products. Am J Public Health 2019;109:960–61.

